# Protective association between recent insecticide-treated bed net use and falciparum malaria in three provinces of the Democratic Republic of the Congo

**DOI:** 10.64898/2026.09.15.26363108

**Authors:** Danielle L. Wiener, Eddy Kieto, Melchior M. Kashamuka, Mvuama Nono, Jonathan J. Juliano, Fernandine Phanzu, Adrien N’Siala, Tommy Nseka, Antoinette K. Tshefu, Albert Kalonji, Tim Sheahan, Joris Likwela, Jonathan B. Parr

## Abstract

Recent research highlights threats to the effectiveness of insecticide-treated bed nets (ITNs) against malaria transmission. This cross-sectional study (N=2819) investigated associations between bed net usage and malaria rapid diagnostic test (RDT) positivity among those presenting with malaria symptoms in three provinces of the Democratic Republic of the Congo (DRC). A log-binomial regression model was used to determine RDT-confirmed malaria prevalence ratios; prior malaria diagnosis and location of care were assessed as modifiers. Using a bed net the prior night was associated with lower risk of malaria diagnosis [PR of 1.39 (95% CI: 1.20, 1.62), with a quantitative bias analysis adjusted result of PR = 1.64]. ITN use reduced risk of malaria diagnoses regardless of prior malaria infection. However, prior diagnosis did modify the relationship between bed net usage and malaria RDT positivity, with prior diagnosis reducing bed net effectiveness. These findings add to existing literature showing the benefits of ITN use in highly malaria-endemic settings like the DRC.

## Introduction

Malaria remains a major public health problem, having caused approximately 249 million cases and 631,000 deaths in 2022.^1^The primary effective interventions against malaria which target the mosquito vector include insecticide-treated bed net (ITN) use and indoor residual spraying (IRS)^1^. For most malaria endemic regions, ITNs are the primary vector control intervention. These interventions, as well as the use of artemisinin-combination therapies (ACTs) for treatment, are threatened by growing resistance to insecticides and antimalarial drugs. ITNs are widely deployed in sub-Saharan Africa, with 282 million distributed to malaria endemic countries in 2022 alone^1^. However, continued investment in effective ITN distribution, proper utilization, and development of new insecticides is needed to achieve malaria control goals.

Evidence regarding the impacts of ITNs can be used to guide allocation of programmatic resources, particularly in high-burden African countries like the Democratic Republic of the Congo (DRC)^2^.

This study aimed to investigate the effectiveness of ITN use against symptomatic malaria in the DRC. The DRC has the second highest number of malaria cases and deaths worldwide, with increasing cases despite an estimated 75% of the population having access to insecticide treated nets (ITNs)^1^. Recent research indicates a lack of equality in ITN access based on wealth and geographic region, and insufficient education and community outreach leading to low usage among adults who own ITNs^3–7^. Several types of netting are used in the DRC, with varying effectiveness against malaria due to durability issues over time and insecticide variety^8–11^.

Increased insecticidal resistance of both *Anopheles gambiae* and *Anopheles funestus*, the primary mosquito vectors transmitting malaria in Kinshasa Province, reduces the protective benefit of ITNs even with adequate ownership and appropriate use^12–14^.

Contemporary evidence regarding the impacts of ITN use on malaria risk in the DRC is needed. Prior studies have focused on high-risk subgroups, including pregnant women or children <5 years of age, on insecticides and resistance, or examined other associations like the impact of education on bed net use^3–13, 15^. The present study provides a broader perspective on the impacts of bed net ownership and use on symptomatic malaria. The presence and magnitude of the relationship between the use of a bed net the night prior to presenting to a health facility with malaria symptoms and RDT positivity was assessed, increasing understanding of the contemporary effectiveness of ITNs in the DRC.

## Methods

### Study Population

This cross-sectional study was a secondary analysis of data collected as part of a previous study of RDT performance and *Plasmodium falciparum* histidine-rich protein 2 and 3 (*pfhrp2/*3) deletions^16^. This study was led by SANRU Asbl to support the DRC’s Programme Nationale de Lutte contre le Paludisme (PNLP), in collaboration with l’Ecole de Santé Publique de l’Université de Kinshasa and the University of North Carolina at Chapel Hill. Details of the parent study have been published. In brief, adults and children presenting with malaria symptoms received RDT diagnosis and treatment according to national guidelines, answered questions as part of a structured questionnaire, and provided dried blood spot samples (DBS) for molecular and serological testing. The project was conducted in December 2017 at 18 health facilities in the Bas Uele, Kinshasa, and Sud Kivu Provinces of the DRC. All participants provided consent or, for minors, assent alongside parental consent. Consenting patients who presented to the health centers with suspected malaria were considered eligible for inclusion. De-identified data were accessed for research purposes on September 11, 2023.

### Variable Measurement

Analysis focused on ITN use and RDT results. The main exposure was self-reported bed net use, defined as whether the participant recalled having used an ITN the night before. It was used as an indicator for likely recent bed net use habits prior to transmission. For validation of behavior patterns, this indicator variable was statistically compared to another variable for the number of times in the past 7 days that the participant slept under a bed net. For parsimony of the model and to avoid reducing statistical power, the binary exposure of prior night usage was chosen over the categorical self-reported ITN usage over 7 nights, which included 8 possible exposure categories (0-7).

Detailed information from the parent study included microscopy results, species of *Plasmodium* detected based on real-time PCR, quantitative real-time PCR (qPCR) targeting the single-copy *P. falciparum* lactate dehydrogenase (*pfldh*) gene, and other biological variables. The RDT used was the World Health Organization- (WHO-) prequalified, SD BIOLINE Malaria Ag P.f. (05FK50, Alere, Waltham, MA) which is an HRP2 only test. Results from RDTs were chosen as the primary outcome over other diagnostic measures as they are able to capture both active and recent infection. HRP2-based RDTs generally detect antigen for a few weeks after parasite clearance.^17^ Primary covariates analyzed included the Provincial Health Department (DPS) [corresponding to the location where the patient reported for care], pregnancy status, age, previous diagnosis of malaria within 6 months prior, and reported biological sex.

The outcome measure was coded to allow for the reference group to be those who did use a bed net. DPS was a categorical variable with three categories (Kinshasa, Sud Kivu, and Bas Uele), using Kinshasa as the reference based on city residents likely having better access to education and bed nets.^4 6 7^ Sex, prior diagnosis, and pregnancy status were all categorical variables, while the quadratic spline functional form was used for age after analysis of options (including AIC and predicted value plots). Knots were placed at 10, 20 and 60, based on a locally weighted scatterplot smoothing (LOWESS) plot.

### Statistical Analysis

The overall association between bed net usage and symptomatic, RDT-confirmed falciparum malaria (primary outcome) was estimated, along with the effect of potential modifying variables (prior malaria diagnosis and DPS). A log-binomial regression model was fitted based on the DAG conceptualization of variables (Supplemental Figure 1), and prevalence ratios (PRs) were produced as a measure of association. PRs for this study represented the prevalence of RDT+ malaria among symptomatic patients presenting for care. The model was adjusted for pregnancy status, age, and prior diagnosis, and an interaction term for prior diagnosis and bed net use was evaluated. Effect measure modification (EMM) and the presence of a joint effect were evaluated based on substantive knowledge, graphical representation, a Wald Chi-Square test, and a modification table comparing expected to observed modification. Width and precision of the 95% confidence intervals (CIs) were considered. Statistical analyses were performed using Statistical Analysis Software (SAS) version 9.4 software.

Sensitivity for detecting *P. falciparum* infection using mRDTs generally varies by season (wet/dry) and ranges from 86% (85% CI: 78-92%) to 94% (95% CI: 92-96), with specificity between 90% (95% CI: 86-92) and 78% (95% CI: 72-83%)^18^. Prior research using the same dataset indicated that RDT performance varied by region in the DRC, and that RDT sensitivity and specificity were 75% and 92%, respectively, compared to PCR^16^. A quantitative bias analysis was conducted to analyze the sensitivity of the primary regression estimate to bias due to RDT mismeasurement. Input of observed data was based on the following numbers: RDT-/No bed net use = 311, RDT+/No bed net use = 265, RDT-/Bed net use = 1418, RDT+/Bed net use = 820. Bed net use was considered the referent group for this analysis. Outcome misclassification bias was measured using Excel Spreadsheet macros based on methodology developed by Lash and colleagues^19^.

## Results

Among 3,627 individuals enrolled in the parent study, this analysis sample consisted of 2819 individuals after exclusion based on missing exposure data (n=813). 58.7% of the sample was female, 42.6% were RDT-positive, and 77.2% were ≤ 34 years old. 80% of analyzed participants used a bed net, 61% had negative mRDT results, and 42% had a previous malaria diagnosis within 6 months. Participants were enrolled in Kinshasa (40%), followed by Sud-Kivu (33%) and Bas-Uele (27%). Females in the analysis sample moderately outweighed the males (59%), and of those eligible, 18% were pregnant. The median age was 20 years (25^th^percentile = 5, 75^th^percentile = 33; range 0-93), with minimal missingness (n=50, 0.02%), showing that this population had an age range skewed towards younger individuals.

Indicator variable accuracy was assessed via cross-tabulation comparison. Of those reporting not having slept under a bed net the night prior to healthcare presentation, 66.3% had used a bed net 0 times in the last week (7 times = 2.1%, 3+ times = 27.9%). Of those who reported they did use a bed net the prior night, 81.9% had slept under a bed net all 7 nights (3+ times = 96.8%, 0 times = 1.1%). These results were considered to have sufficiently concordant behavior patterns.

The final regression model highlighted the protective nature of bed net usage against RDT-confirmed falciparum malaria. An overall prevalence ratio (PR) of 1.39 (95% CI: 1.20, 1.62) indicates that participants who did not report bed net use were 1.39 times more likely to have a malaria diagnosis than participants who reported bed net use, after adjusting for prior diagnosis, age, and pregnancy status (Table 2). Results of the QBA using information on sensitivity and specificity for this dataset gave a corrected PR = 1.64, which was just outside the bounds of the original effect estimate CI. As such, bias of the PR estimates caused by RDT measurement error was likely to be minimal. As seen in Table 2, the association generally did not differ significantly by location of care. Benefits of recent ITN use were more pronounced in Kinshasa, where the PR was 1.39 (95% CI: 1.07, 1.82), compared to PRs of 0.85 (95% CI: 0.61, 1.18) in Sud-Kivu and 1.15 (1.00, 1.31) in Bas-Uele.

**Table 1.**
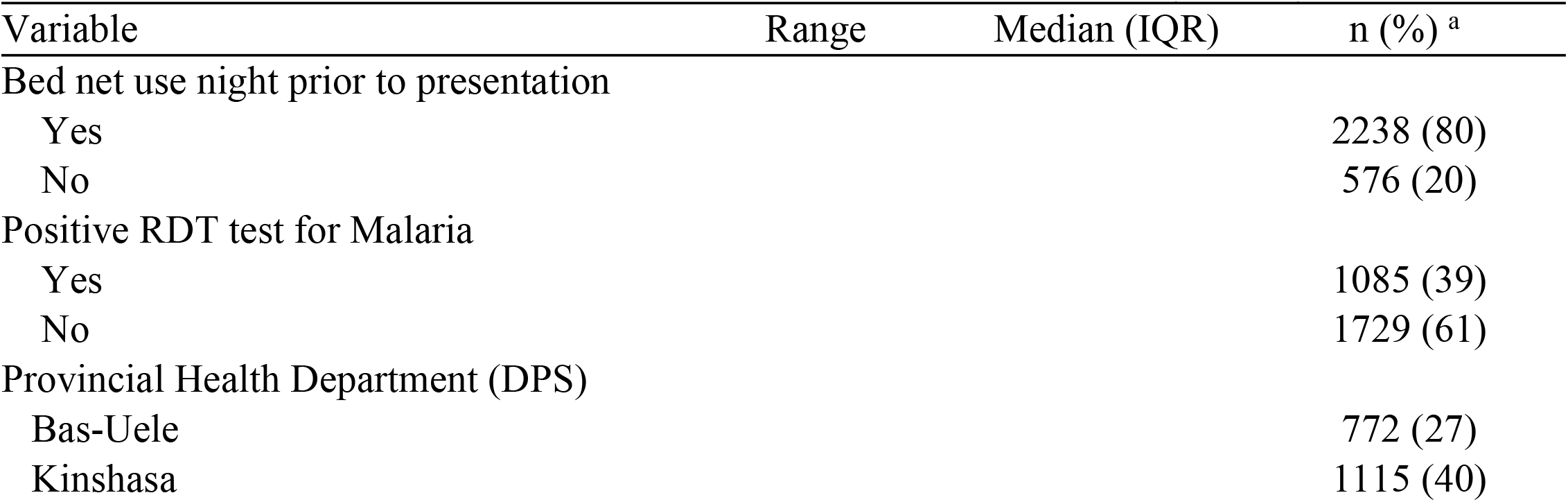
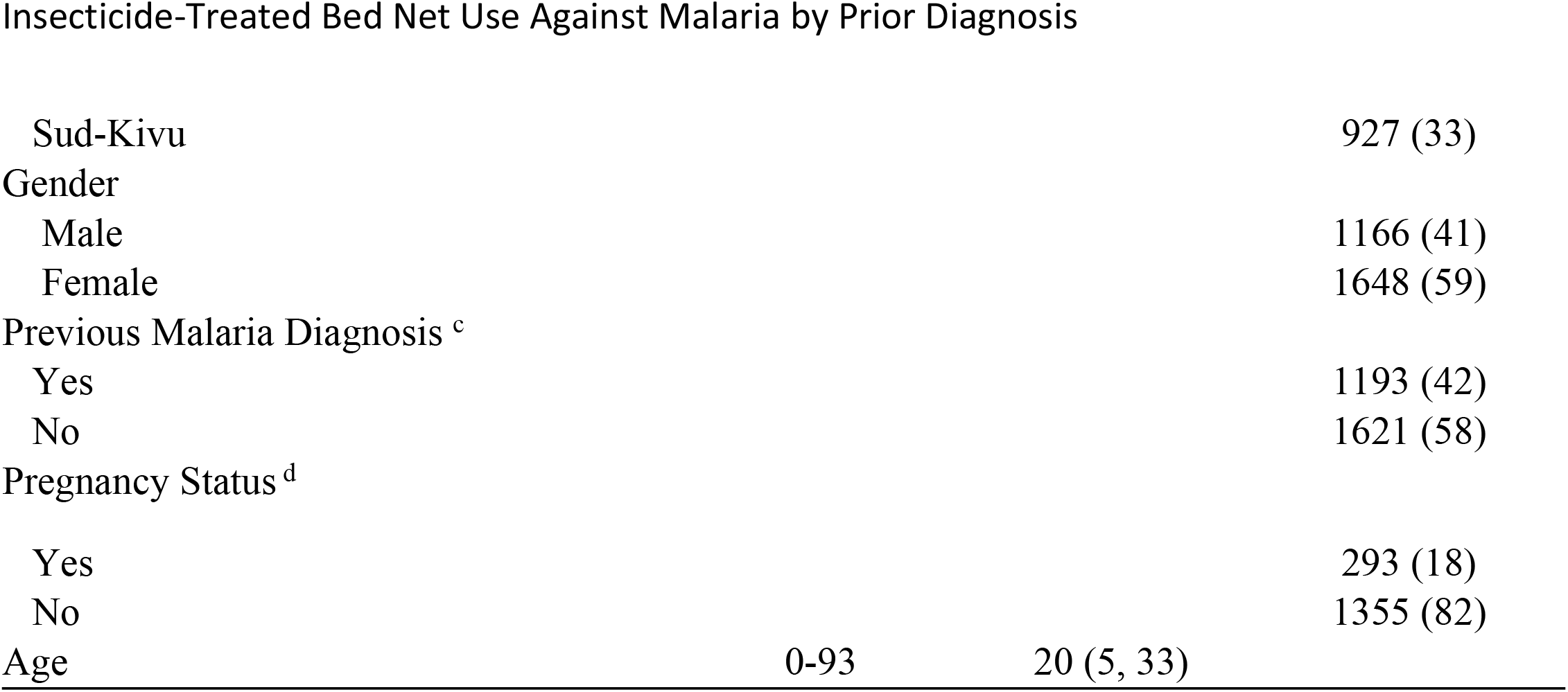
Descriptive Characteristics of DRC Cross-sectional Study Sample (N=2819)

**Table 2.** Adjusted Modification of the Effect of No Bednet Use on Malaria Results by Previous Diagnosis ^e f g h i^.

| Prior RDT-positive<br>*P.falciparum*<br>malaria diagnosis* | No Bed Net Use Night Prior to Presenting |  | Bed Net Use Night Prior to Presenting |  | PRs (95%CI) for no<br>bed net use within<br>strata of previous<br>diagnosis |
| --- | --- | --- | --- | --- | --- |
|  | N with/without Malaria | PR (95% CI) | N with/without Malaria | PR (95% CI) |  |
| **Overall** |  |  |  |  |  |
| No Prior Diagnosis | 141/207 | 1.39 (1.20, 1.62)<br>p <.0001 | 369/904 | 1.00 | 1.39 (1.20, 1.62)<br>p <.0001 |
| Prior Diagnosis | 124/104 | 1.78 (1.55, 2.04)<br>p <.0001 | 451/514 | 1.56 (1.40, 1.74)<br>p <.0001 | 1.27 (1.01, 1.50) |
| **Kinshasa** |  |  |  |  |  |
| No Prior Diagnosis | 53/96 | 1.39 (1.07, 1.82) | 136/410 | 1.00 | 1.39 (1.07, 1.82) |
| Prior Diagnosis | 29/35 | 1.73 (1.27, 2.34) | 118/238 | 1.32 (1.07, 1.63) | 1.24 (.88, 1.75) |
| **Sud-Kivu** |  |  |  |  |  |
| No Prior Diagnosis | 31/88 | .85 (.61, 1.18) | 147/388 | 1.00 | .85 (.61, 1.18) |
| Prior Diagnosis | 24/34 | 1.35 (1.00, 1.82) | 82/133 | 1.14 (.93, 1.41) | 1.59 (1.06, 2.38) |
| **Bas-Uele** |  |  |  |  |  |
| No Prior Diagnosis | 57/23 | 1.15 (1.00, 1.31) | 86/106 | 1.00 | 1.15 (1.00, 1.31) |
| Prior Diagnosis | 71/35 | 1.10 (.98, 1.24) | 251/143 | 1.10 (1.00, 1.21) | .96 (.85, 1.09) |

We also observed evidence to support modification of the relationship between bed net usage and malaria RDT positivity by prior diagnosis, consistent with expectation. Crude estimates of modification showed PRs of 1.39 (95% CI: 1.20, 1.63; p<.0001) and 1.16 (95% CI: 1.02, 1.33, p=.0297) for individuals with no prior malaria diagnosis in the preceding 6 months and for individuals with prior diagnosis in this time frame, respectively. After adjustment, malaria diagnoses remained more common without bed net use among those with prior infection [PR = 1.27 (95% CI: 1.01, 1.50)] (Table 2). For individuals without prior diagnosis, the adjusted PR was equivalent to the crude PR estimate. There was evidence to support that prior exposure to malaria impacts the protectiveness of bed net use (Wald statistic = 4.09; p = 0.04; α = 0.05).

Joint effects were assessed to determine whether the observed PR for the doubly exposed (no bed net use and with prior diagnosis) group differed from the expected PR on the multiplicative scale. An overall interaction contrast ratio (ICR) of −0.17 indicated that there was a departure from perfect multiplicity. Considering this, prior diagnosis seemed to have an antagonistic effect on the relationship by reducing the positive benefit of using a bed net. For example, among those with a prior diagnosis in Sud-Kivu, participants who did not report bed net use were 1.59 (95% CI: 1.06, 2.38) times more likely to have a positive malaria test. In comparison, those in the same region but in the no prior infection stratum only had a PR of 0.85 (95% CI: 0.61, 1.18). This shows that the risk of malaria RDT positivity without ITN usage was increased amongst a population with previous RDT+ infections.

## Discussion

Our findings underscore the value of ITNs as an important intervention for reducing *P. falciparum* malaria cases in the DRC. Because their effect can change based on transmission intensity, evolution of insecticide resistance, behavior patterns, age of the net, and other factors, iterative evaluation of ITN effectiveness in real populations is needed to help inform control programs. Results of this study show that ITN usage has a protective relationship against malaria diagnosis. The QBA indicated that there may be mild bias towards the null for these PR estimates based on prior sensitivity and specificity research using this dataset. As such, it is likely that bed net use offers an even greater protectiveness than is suggested by our estimates.

The protectiveness of ITN usage is modified by history of prior diagnosis. As expected, prior diagnosis reduces the protective benefit of ITNs. These results are in line with literature suggesting that re-occurrence or re-presentation for healthcare is not uncommon due to continued habitation in endemic areas, ongoing clearance of an initial infection, or re-infection from a different mosquito^19^. Despite this modification, use of a bed net the night before was associated with reduced risk of symptomatic malaria among both individuals with and without prior malaria diagnosis.

This pattern was observed within all regions sampled, except for those in the no prior malaria diagnosis group in Sud-Kivu and the prior diagnosis group in Bas-Uele. For Bas-Uele, the Hessian convergence criterion was not met due to small sample size and thus those results may not accurately portray the true PRs. The PR estimate below the null in Sud-Kivu may indicate that there are other unmeasured factors affecting the effectiveness of the intervention in that area. Sud-Kivu may be an area where more targeted interventions are required among those without a prior diagnosis of malaria, as the current bed net usage does not appear to be as effective (PR = .85, 95% CI: .61, 1.18). Acknowledging that the CIs overlap, and that the result could be random chance within the sample, it is also possible that study site characteristics or the location of Sud-Kivu next to two large water-resource reservoirs (Lakes Kivu and Tanganyika) may play a role. For example, wind around large water bodies such as these has been shown to play a considerable role in *Anopheles* mosquito population dynamics^20^. The difference may also be due to older or less effective netting in circulation, the presence of historical sociopolitical inequality in less urban areas, or insufficient education on bed net use^3–14, 21–24^. Alternatively, a larger sample size of participants within these provinces may resolve the discrepancy. Regardless, recent malaria diagnosis clearly influences the efficacy of malaria ITN interventions.

This study is not without limitations. First, selection bias is possible as data was collected only for patients who presented at health facilities. This may exclude those unable to present for care for various reasons, therefore limiting generalizability to symptomatic individuals without challenges to accessing healthcare centers. It is possible that these challenges overlap with reduced access to ITNs or knowledge of ITN usage, which could lead to differences in malaria incidence among this population. Second, RDT measurement error is expected and could influence our results. Misclassification of the outcome was addressed through QBA, which gave a corrected main effect estimate just beyond the CI of the original and indicated that bias from this source was likely mildly towards the null. Third, unintended biases inherent to the original study or current study design, missing variables, and sparsity are possible. Fourth, we do not have data about the specific ITN types available within households, and therefore cannot comment on the probability of insecticide resistance in a given region. Finally, our analysis of cross-sectional observational data demonstrates important associations but cannot establish causality. Randomized trials of different ITN types, or distribution and educational approaches are needed to determine the true impacts of contemporary ITN strategies in the DRC.

In summary, this study shows a protective association between sleeping under a bed net and RDT-confirmed, symptomatic falciparum malaria. We explore nuances of modification by past RDT diagnosis and geographical region, with results showing protection across most comparisons. We are unable to determine reasons for provincial differences in the benefits of ITN use, but possible explanations include insufficient education and community outreach, varying quality of bed nets, low usage levels, and insecticidal resistance as contributing to increased malaria risk^3–14^. Sustained efforts to overcome these barriers are needed to maintain the protective benefits of ITNs in the DRC.

## List of Abbreviations

ITNs: insecticide-treated bed nets
RDT: rapid diagnostic test
DRC: Democratic Republic of the Congo
QBA: Quantitative bias analysis
PR: prevalence ratio
IRS: indoor residual spraying
ACTs: artemisinin-combination therapies
DBS: dried blood spot samples
qPCR: quantitative real-time PCR
WHO: World Health Organization
DPS: Provincial Health Department
LOWESS plot: locally weighted scatterplot smoothing
NA: Not Applicable
DAG: Directed Acyclic Graph
EMM: Effect measure modification
CIs: confidence intervals
SAS: Statistical Analysis Software
mRDT: malaria rapid diagnostic test ()

## Declarations

### Consent for publication

All participants provided consent or, for minors, assent alongside parental consent.

### Availability of data and materials

The datasets used and analyzed during the current study will be made available through the UNC Dataverse (ID pending).

## Authors’ contributions

DLW, JJJ, TS, and JBP conceptualized the study. DLW wrote the first draft and conducted all analyses. EK, MMK, MN, FP, AN, TN, AKT, AK, JL, and JBP played key roles in data and sample collection and analysis in the parent study. All authors reviewed the final manuscript.

## Acknowledgements

Not Applicable

---

**Supplemental Figure 1.**
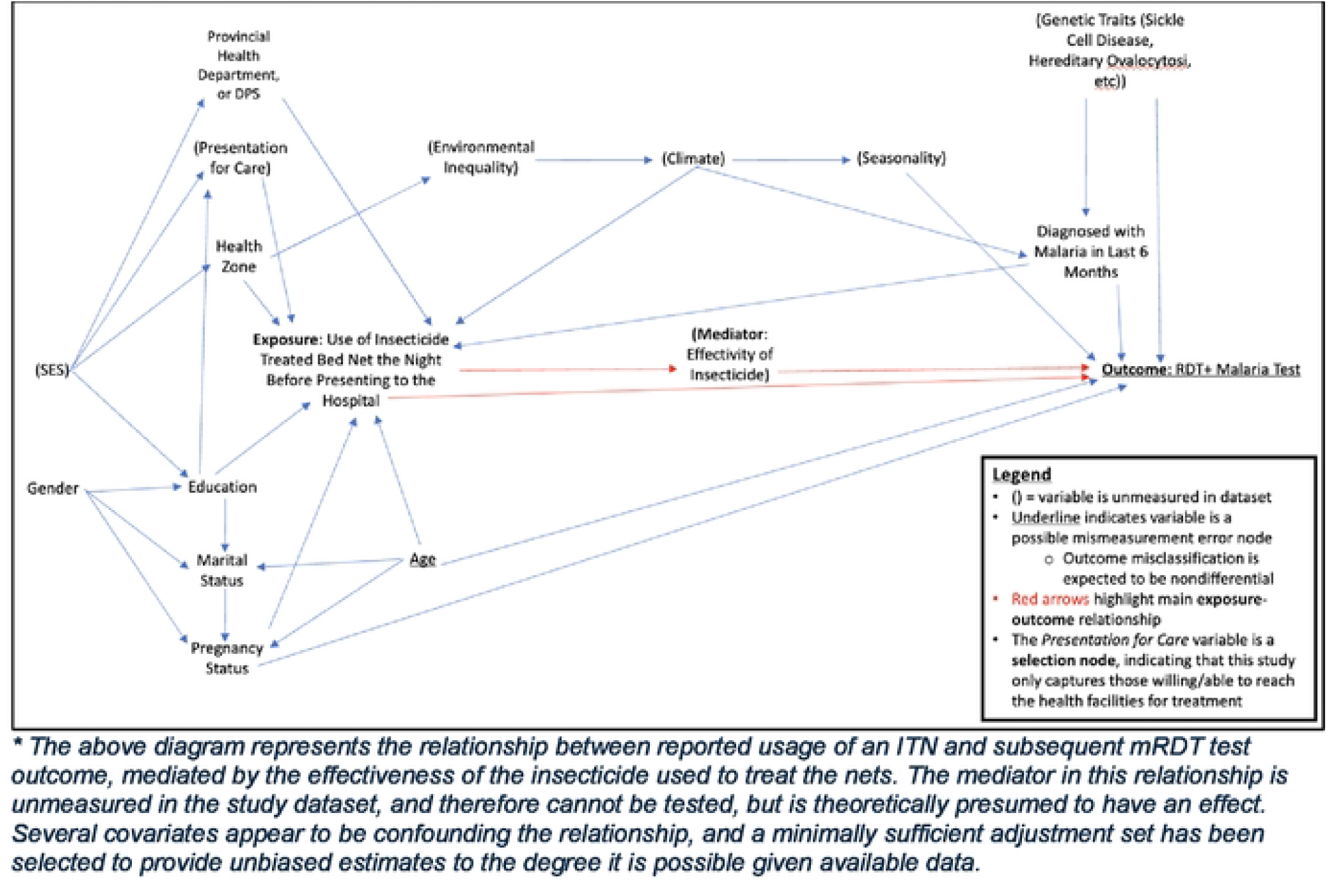
Directed Acyclic Graph (DAG) of the Relationship between Bed Net Usage and Malaria RDT Test Outcome. ^★^*The above diagramrepresents the relationship between reported usage of an ITN and subsequent mRDT test outcome, mediated by the effectiveness of the insecticide used to treat the nets. The mediator in this relationship is unmeasured in the study dataset, and therefore cannot be tested, but is theoretically presumed to have an effect. Several covariates appear to be confounding the relationship, and a minimally sufficient adjustment set has been selected to provide unbiased estimates to the degree it is possible given available data*.

## Footnotes

a Percent of non-missing observations. No variables had missing values expect for age (n= 50).

b Percent of all observations

c Within 6 months prior to presenting to health center for this study.

d 1166 of individuals in sample were not applicable (NA) for pregnancy status analysis in this sample due to being male. Numbers shown are only for female participants.

e Prior Diagnosis indicates participant was RDT-positive for falciparum malaria within 6 months before presenting to the healthcare center for this event.

f Wald test for interaction value; Overall = 4.09, p = 0.04; Overall ICR = −.17

g Adjusted for age, pregnancy status, prior diagnosis; stratified by provincial health department (DPS).

h Due to smaller sample size numbers from Bas-Uele, the relative Hessian convergence criterion of 0.118 was greater than the limit of 0.0001.

i Prevalence Ratios (PRs) are given for the association of Bednet Use and mRDT results, stratified by region. Both singly exposed and the doubly subgroups with each strata are compared to the reference group of doubly unexposed group within the same strata.

